# Use of excess mortality associated with the COVID-19 epidemic as an epidemiological surveillance strategy - preliminary results of the evaluation of six Brazilian capitals

**DOI:** 10.1101/2020.05.08.20093617

**Authors:** André Ricardo Ribas Freitas, Nicole Montenegro de Medeiros, Livia Carla Vinhal Frutuoso, Otto Albuquerque Beckedorff, Lucas Mariscal Alves de Martin, Marcela Montenegro de Medeiros Coelho, Giovanna Gimenez Souza de Freitas, Daniele Rocha Queiróz Lemos, Luciano Pamplona de Góes Cavalcanti

## Abstract

In early 2020, the World Health Organization (WHO) recognized the pandemic situation of the new coronavirus (severe acute respiratory syndrome coronavirus 2, SARS-CoV-2), which causes Coronavirus Disease-2019 (COVID-19). In Brazil by the end of April 2020, another 110 thousand cases and 5,000 deaths had been confirmed. The scarcity of laboratory resources and overload of the care network, added to the broad clinical spectrum of the disease, can make it difficult to capture all mortality from this disease through epidemiological surveillance based on individual notification of cases. The aim of this study was to evaluate the excess of deaths in Brazilian capitals with the highest incidence of COVID-19, as a way of validating the method, we also evaluated a capital with low incidence.

We assessed weekly mortality from all causes during the year 2020, up to the epidemiological week 17, compared with the previous year. The data were obtained through the National Civil Registry Information Center (CNIRC, acronym in Portuguese). We estimate the expected mortality and the 95% confidence interval by projecting the observed mortality in 2019 for the population of 2020.

In the five capitals with the highest incidences it was possible to identify excess deaths in the pandemic period, the age group most affected were those over 60 years old, 31% of the excess deaths occurred in the population between 20 and 59 years old. There was a strong correlation (r = 0.94) between the excess of deaths in each city and the number of deaths confirmed by epidemiological surveillance. There was no excess of deaths in the capital with the lowest incidence, nor among the population under 20 years old. We estimate that epidemiological surveillance managed to capture only 52% of all mortality associated with the COVID-19 pandemic in the cities studied.

Considering the simplicity of the method, its low cost and reliability for assessing the real burden of the disease, we believe that the assessment of excess mortality associated with the COVID-19 pandemic should be widely used as a complementary tool to regular epidemiological surveillance and its use should be encouraged by WHO.

## INTRODUCTION

In December 2019, an increase in hospitalizations for pneumonia of unknown etiology was observed in Wuhan, Hubei, China. Initially the disease was associated with exposure in a seafood and wild animal market, but then transmission from person to person was confirmed. On January 7, 2020 a new coronavirus was isolated and identified as the cause of the pneumonia outbreak, January 13 was confirmed the first case in Thailand, on January 19, 2020 the first case was confirmed in Korea, two cases in Beijing and one case in Guangdong (China)[1]. On February 11, the World Health Organization (WHO) announced the name of Coronavirus Disease-2019 (COVID-19), having as main symptoms in the acute phase of the disease fever, cough, myalgia and dyspnea. On the same day, the International Committee on Taxonomy of Viruses (ICTV) named this new coronavirus as severe acute respiratory syndrome coronavirus 2 (SARS-CoV-2). WHO technical visit carried out between 16 and 24 February estimated the R0 basic number of reproduction cases of the then new coronavirus initially between 2-2.5[2]. Even before WHO recognized that the occurrence of the coronavirus was a pandemic (March 11), the virus had already spread to many countries. By the end of April, more than 3.6 million cases and 251.5 thousand deaths had been registered on the five continents. [2]. In Brazil, the first case of COVID-19 was confirmed on February 26, 2020. After 15 days, sustained transmission was declared and on March 17, the first death was confirmed. By the end of April 2020, more than 110 thousand cases and 5,000 deaths had been confirmed, with a mortality coefficient of 20 deaths per 1 million inhabitants, with Brazil among the 10 countries with the highest number of deaths.[3].

Despite the high number of confirmed cases and deaths in Brazil to date, and efforts to diagnose suspected cases, it is believed that testing is far from necessary. The COVID-19 testing laboratory network was established gradually, first centralized in three units of National Influenza Center (NIC) and later included private laboratories and the state public health laboratory network [4]. The WHO recommendation on March 16, of mass testing for COVID-19 resulted in a shortage of tests on a worldwide scale that, added to the increase in cases in the country, compromised the testing capacity in Brazil, not only in the public network, but also in the private health network[4], this may be contributing to the underestimation of the real magnitude of the disease in the country, the same may be happening worldwide.

The broad clinical spectrum of the disease, which can hinder clinical diagnosis, added to the scarcity of laboratory resources and the overload of the health care network, will not allow epidemiological surveillance systems to detect all cases and deaths. A reliable assessment of the burden of COVID-19 is essential so that health managers and country governments can better decide on the magnitude and duration of non-pharmacological interventions to control the disease. The objective of this study was to analyze the excess of deaths and the mortality coefficient due to the current SARS-CoV-2 pandemic, in the capitals with the highest number of cases recorded in routine surveillance systems, in Brazil.

## METHODS

This is a study based on official public data in which we analyzed the time series, cases, deaths due to infection by COVID-19 and the excess of deaths in the first months of the pandemic in six Brazilian capitals.

### Study locations

For the present study, the five Brazilian capitals with the highest absolute numbers of confirmed cases of COVID-19, Fortaleza, São Paulo, Rio de Janeiro, Manaus and Recife were selected (Figure - 1). To validate the method, the capital Porto Alegre was also studied, which has one of the lowest incidence rates of COVID-19 and which recently conducted a serological survey proving a low seroprevalence rate.[5]

**Figure 1.**
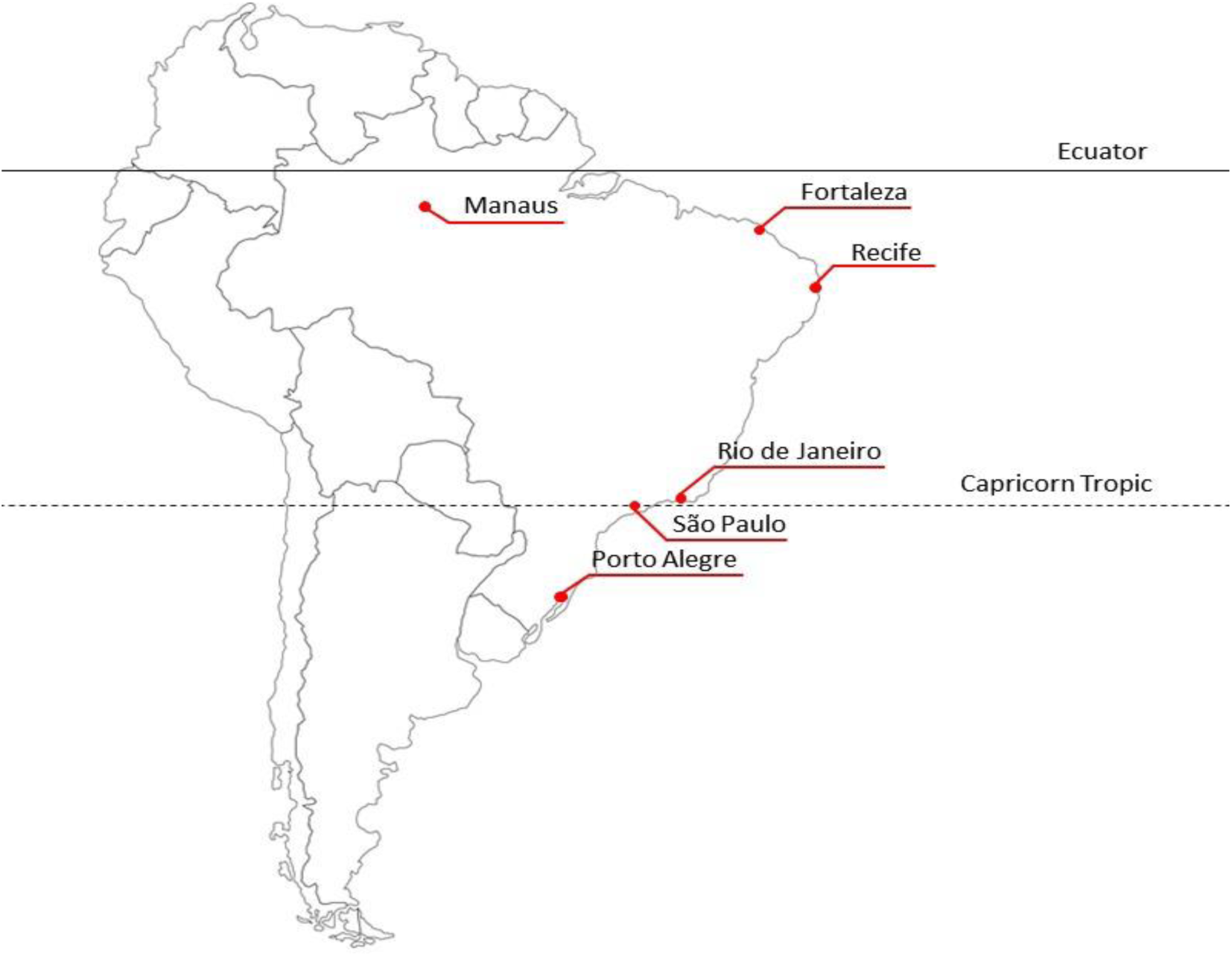
Location of the six capitals studied.

### Data sources

Mortality data were collected on the website of the National Civil Registry Information Center (CNIRC, acronym in Portuguese)[6], that made available information on deaths registered in notary offices across the country in the years 2019 and 2020. In Brazil, notary offices have a legal period of 14 days to enter the data contained in death certificates signed by the attending physician in the CNIRC. Deaths due to COVID-19, pneumonia, severe acute respiratory syndrome, septicemia, undetermined causes and other deaths were included in the analysis, the criteria used are presented in Annex I. Deaths from external causes were excluded. In Brazilian law, the definition of the cause of death in the death certificate is preferably made by the attending physician and there is no obligation for the diagnostic hypothesis to have laboratory confirmation. Therefore, we group deaths as respiratory causes (COVID-19, pneumonia and severe acute respiratory syndrome) and all causes (including previous and more septicemia, other deaths and undetermined causes).

Data from confirmed cases and deaths by COVID-19 were collected in the repository of the Brazil project, which performs the extraction from official sources following a preestablished pattern [7] deaths are classified according to the date of confirmation of the case in the Notification System for Notifiable Diseases (SINAN acronym in Portuguese) and are available for wide consultation [8].

To obtain the COVID-19 mortality data confirmed in SINAN we search the death dates directly in the databases and reports of the State and Municipal Health Secretariats [9–11]. It was not possible to obtain data by symptom onset date for the municipalities of Rio de Janeiro and Manaus. In order to make it possible to compare the temporal quality of the information, all data were obtained on the same day (May 2, 2020).

Demographic data were collected on the Brazilian Institute of Geography and Statistics (IBGE, acronym in Portuguese) website, which estimates the population of Brazilian capitals by age group on a quarterly basis through the Continuous National Household Sample Survey (PNADc, acronym in Portuguese)[12]. For the first quarter of 2019 the estimated population for the last quarter of 2018 was used and for 2020 the estimated population for the last quarter of 2019 was used.

### Data analysis

We assessed weekly mortality from all causes during the year 2020, up to the epidemiological week 17, compared with the previous year. We calculated the expected mortality for 2020 and the 95% confidence interval, projecting the 2019 mortality rate for the 2020 population.

#### Definition of the period with excess mortality

The period with excess mortality was defined as starting in the first sequence of two consecutive weeks with a death toll above the upper limit of 95% confidence interval and ending in the first of two consecutive weeks with a death toll below this limit[13].

As the mortality data can be entered up to 14 days after death, we evaluated the correlation between the cases officially confirmed by the Health Departments and the excess of deaths until the epidemiological week 16, then we constructed a linear regression.

The mortality rate by age group was calculated for the period with excess mortality. The expected number of deaths by age group was calculated by projecting the 2019 death rate by age group for the 2020 population. Excess mortality by age group was calculated as the number of deaths observed minus the expected number of deaths. The ratio of mortality rates was calculated by dividing the mortality rate by age group in 2020 during the period with excess mortality, by the mortality rate observed in 2019.

Statistical analyzes were performed using the STATA 9.2 program and the results presented in graphs and tables built in the Microsoft® Excel® program..

## RESULTS

Table 1 shows the population and the epidemiological situation of the cities studied. We also present an excess of deaths from all causes during the period classified as excess mortality. We can observe different incidence rates per 100,000 inhabitants, the lowest being 3.3 in Porto Alegre and the highest 229.2 in Recife. The case fatality rate varied between 3.3% in Porto Alegre, the highest rate was Manaus, 10.2%.

**Table 1.** Population, cases of COVID-19, deaths confirmed by COVID-19 and excess mortality in six Brazilian capitals (data collected until May 2, 2020)

|  | Population<br>(x1,000) | COVID-19<br>Confirmed<br>cases<sup>1</sup> | COVID-19<br>Confirmed<br>deaths<sup>1</sup> | Excess<br>deaths | Confirmed<br>deaths/<br>Excess | IR<sup>2</sup> | CFR<sup>3</sup> |
| --- | --- | --- | --- | --- | --- | --- | --- |
| **Fortaleza** | 2,667 | 6,082 | 413 | 193 | 47% | 228.0 | 6.8 |
| **Manaus** | 2,197 | 3,491 | 357 | 1355 | 380% | 158.9 | 10.2 |
| **Recife** | 1,649 | 3,780 | 250 | 444 | 178% | 229.2 | 6.6 |
| **Rio de Janeiro** | 6,729 | 6,189 | 574 | 1350 | 234% | 92.0 | 9.3 |
| **São Paulo** | 12,278 | 19,085 | 1607 | 2002 | 125% | 155.4 | 8.4 |
| **Porto Alegre** | 1,485 | 455 | 15 | - |  | 30.6 | 3.3 |

<sup>1</sup>Source (SINAN, official health surveillance system, until may 2, 2020)
<sup>2</sup>IR: Incidence Rate (cases/100,000 population)
<sup>3</sup>CFR: case fatality rate (deaths/100 cases)

Figure 2 shows that, with the exception of Porto Alegre, in the 5 cities with the highest incidence of COVID-19, excess mortality was identified. In the cities of São Paulo and Rio de Janeiro there was an increase above 95% of the confidence interval in general mortality from the epidemiological week 11 when compared to the previous year. In these cities, the first cases confirmed by laboratory diagnosis occurred respectively in epidemiological weeks 12 and 13. These findings suggest that there may have been underreporting of cases and deaths by COVID-19 for epidemiological surveillance, but that it can be identified by the syndromic surveillance of excess deaths. In the cities of Recife, Fortaleza and Manaus the excess of deaths can be identified from week 14. In Manaus, the risk of dying in SE 17 from respiratory disease was 11 times greater than in the same period of the previous year, considering all causes of deaths the increase was 4 times. Assuming that week 17 probably still does not include all deaths, these numbers are expected to be even higher. In figure 3-A we can see the strong Pearson correlation (r = 0.95) between the number of confirmed deaths in SINAN and the excess of deaths up to epidemiological week 16, in the six cities studied. The slope of the regression suggests that there was an excess of deaths 2.5 times greater than the official number of deaths reported in week 16 (R^2^=0.91; p<0.0001).

**Figure 2.**
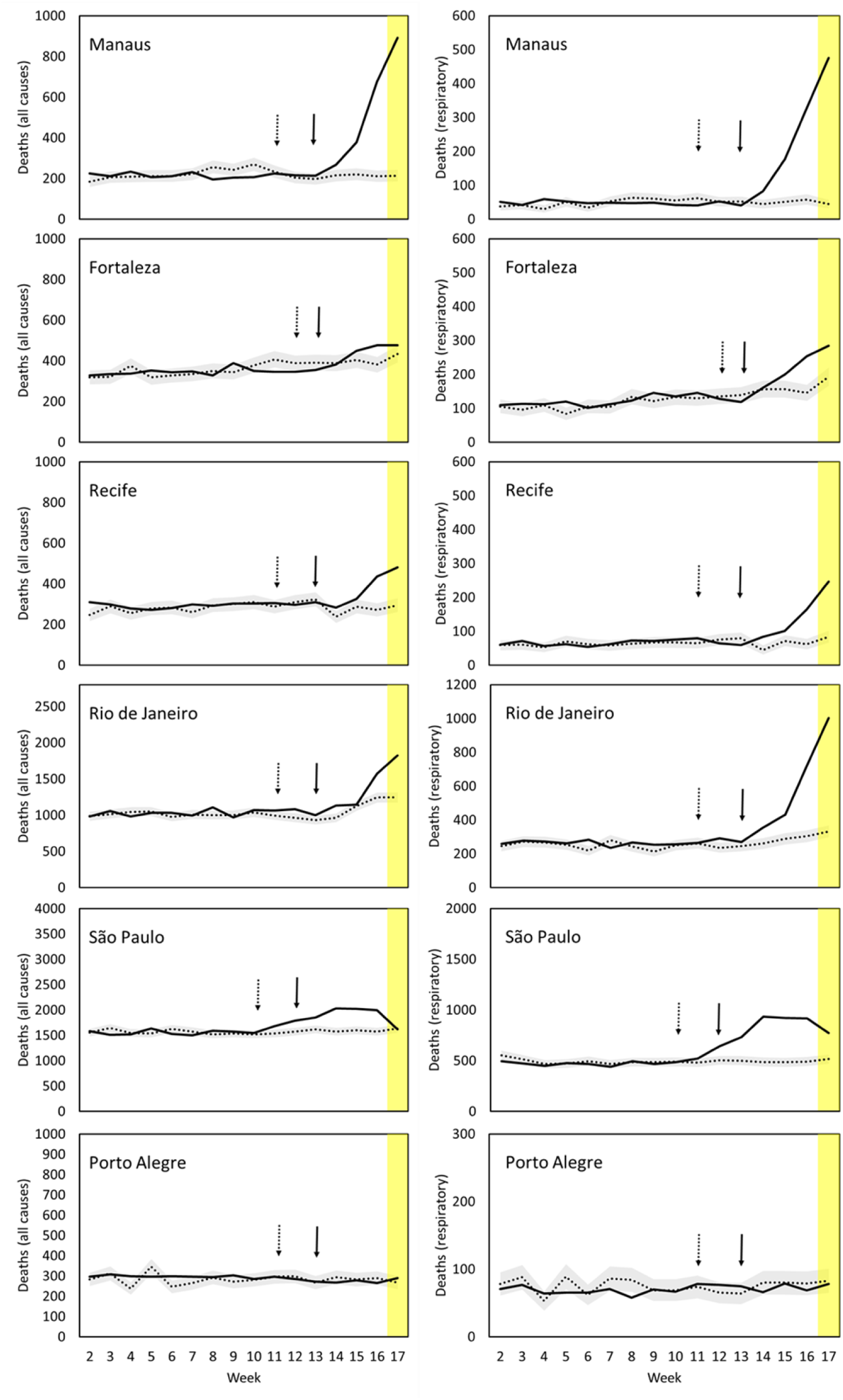
Weekly mortality observed in 2020 (black line), expected (dotted) 95% confidence interval (gray area) for respiratory causes (left) and for all causes (right) by city. The dotted arrow indicates the date of confirmation of the first case and the black arrow indicates the date of the first confirmed death in the city. The yellow area represents the period when mortality data is still incomplete.

**Figure 3.**
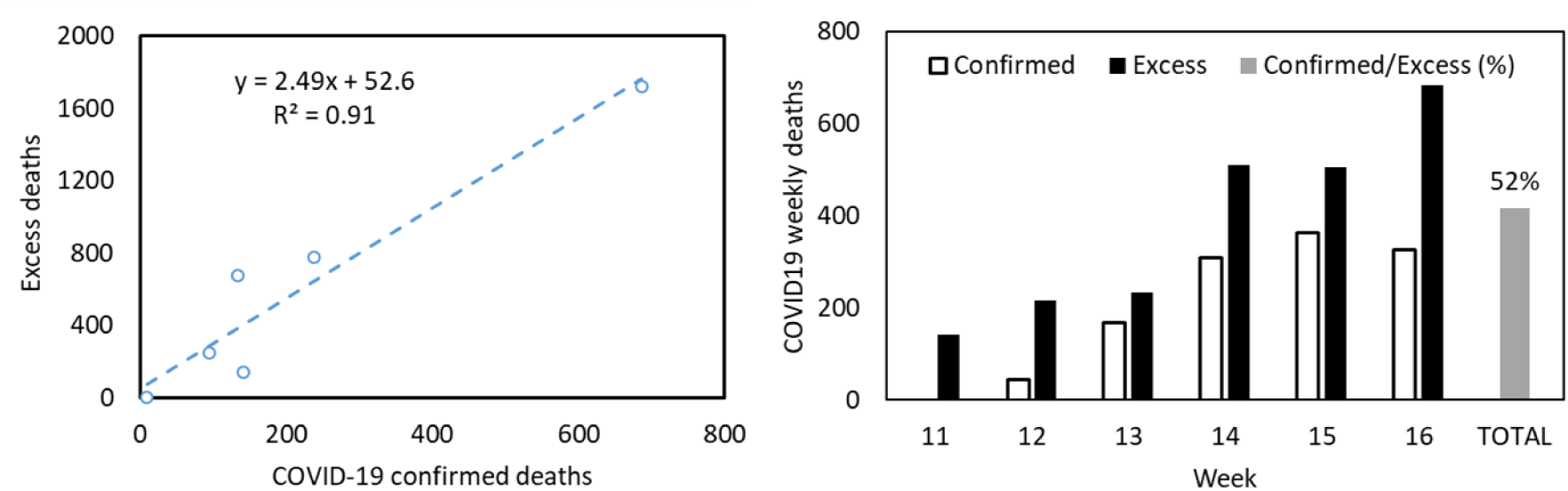
On the left correlation between mortality officially confirmed by COVID-19 in the cities studied and the estimated excess mortality, the equation suggests that for each notified patient there were 2.5 excess deaths by week 16. On the right, deaths officially confirmed by COVID-19 and excess deaths per epidemiological week. Data for the cities of São Paulo, Recife and Fortaleza that provide data according to the date of death.

In figure 3-B, made from the data of the municipalities where it was possible to collect mortality data from COVID-19 based on the date of death (São Paulo, Fortaleza and Recife), we can verify that the mortality confirmed by COVID-19 corresponds to 52% of all excess deaths.

Table 2 shows the mortality rate by age group for all causes. It is possible to verify that the age group most affected was over 60 years old with 3,801 excess deaths from all causes. There was also an excess of deaths in the population aged between 20 and 59 years representing 31% of the total excess mortality from all causes (1,679 deaths).

**Table 2.** Excess mortality due to all causes during the beginning of the COVID-19 pandemic in the five Brazilian capitals with the highest incidence of the disease (data referring to week 17, data collected until May 2, 2020).

|  |  | Mort rate<br>2019 | Mort rate<br>2020 | Rate ratio<br>2020/2019 | Confidence<br>interval (95%) | Deaths<br>2019 | Deaths<br>2020 | Excess<br>deaths | p |
| --- | --- | --- | --- | --- | --- | --- | --- | --- | --- |
| **Fortaleza** | <20 years | 12,4 | 11,3 | 0.92 | (0.66 - 0.27) | 82 | 77 | -5 | 0.5810 |
|  | 21 - 59 years | 16,4 | 21,1 | 1.29 | (1.09 - 1.52) | 260 | 328 | 68 | 0.0023 |
|  | >60 years | 214,9 | 228,4 | 1.06 | (0.97 - 1.17) | 868 | 998 | 130 | 0.1884 |
|  | **Total** |  |  |  |  |  |  | 193 |  |
| **Manaus** | <20 years | 18,4 | 15,7 | 0.85 | (0.65 - 1.11) | 123 | 111 | -12 | 0.2270 |
|  | 22 - 59 years | 18,5 | 45,2 | 2.44 | (2.10 - 2.86) | 233 | 578 | 345 | <0.0001 |
|  | >60 years | 218,8 | 718,4 | 3.28 | (2.97 - 3.64) | 501 | 1523 | 1022 | <0.0001 |
|  | **Total** |  |  |  |  |  |  | 1355 |  |
| **Recife** | <20 years | 16,0 | 16,0 | 1.00 | (0.70 - 1.43) | 70 | 62 | -8 | 0.9886 |
|  | 23 - 59 years | 24,0 | 33,5 | 1.40 | (1.17 - 1.66) | 224 | 321 | 97 | <0.0001 |
|  | >60 years | 291,5 | 378,1 | 1.30 | (1.18 - 1.42) | 787 | 1142 | 355 | <0.0001 |
|  | **Total** |  |  |  |  |  |  | 444 |  |
| **Rio de Janeiro** | <20 years | 16,9 | 15,4 | 0.91 | (0.76 - 1.10) | 250 | 218 | -32 | 0.3333 |
|  | 24 - 59 years | 35,7 | 48,6 | 1.36 | (1.27 - 1.46) | 1362 | 1842 | 480 | <0.0001 |
|  | >60 years | 410,8 | 437 | 1.06 | (1.03 - 1.10) | 5776 | 6678 | 902 | 0.0003 |
|  | **Total** |  |  |  |  |  |  | 1350 |  |
| **São Paulo** | <20 years | 17,6 | 15,6 | 0,89 | (0.78- 1.01) | 537 | 458 | -79 | 0.0562 |
|  | 25 - 59 years | 32,6 | 41,8 | 1,28 | (1.21 - 1.35) | 2271 | 2960 | 689 | <0.0001 |
|  | >60 years | 368,4 | 418,2 | 1,14 | (1.10 - 1.17) | 8043 | 9435 | 1392 | <0.0001 |
|  | **Total** |  |  |  |  |  |  | 2002 |  |

## DISCUSSION

We observed that there was an excess of 5,350 deaths from all causes during the first weeks of the epidemic in the 5 Brazilian cities most affected by COVID-19. This increase was greater in cities with higher incidences, 31% of this excess of deaths occurred in the population between 20 and 59 years old. These findings are compatible with what has been observed in clinical studies and in the bulletins of Brazilian epidemiological surveillance. There was no excess of deaths among children under 20 years of age, nor an increase in overall mortality in the city of Porto Alegre, where the incidence of COVID-19, until the moment of the analysis, was lower. These aspects reinforce the consistency of these findings.

The number of deaths that occurred until the epidemiological week 16 confirmed by the official epidemiological surveillance system corresponded to 52% of the excess deaths that occurred in the same period. Considering that the data were collected 14 days after the end of week 16, we believe that almost all investigations of deaths should have been closed. Therefore, we can estimate that the epidemiological surveillance system only 52% of the total mortality burden of this disease. This is not an unexpected finding, since other viruses such as influenza and chikungunya are also characterized by a mortality burden that is often greater than that measured through the etiological investigation of each patient. Mortality associated with these viruses can be better assessed by calculating the excess mortality that occurs in periods of epidemic [13,14]. Using a similar methodology, it is possible to estimate that in New York City the excess of deaths in the first four weeks was 12547 and, however, conventional epidemiological surveillance was able to identify 7186 deaths from COVID-19, 57% of the excess deaths in the same period[15]. In Portugal the excess of mortality that occurred between March 1 and April 22 was 3 to 5 times higher than that explained by the deaths reported by COVID-19 officially reported [16]. This underestimation of mortality through surveillance systems can occur as a result of several factors, part of which is explained by failure to report, another part of the difficulty in identifying cases that present in atypical ways and the late complications of the disease. The underestimation of mortality by surveillance systems can occur as a result of several associated factors, part of which is explained by failure in the notification or laboratory investigation of patients who would have a case definition criterion, another part is a consequence of the difficulty in identifying the cases that present atypical clinical forms or deaths from late complications of the disease. In fact, a still unknown proportion of patients with COVID-19 have serious and potentially fatal cardiovascular complications such as acute coronary syndrome, myocarditis, arrhythmias and cardiogenic shock, these cases may not be identified by attending physicians in routine situations. [17].

All-cause mortality exceeded the upper limit of the confidence interval (95%) a week before the first laboratory-confirmed death in the cities of São Paulo and Rio de Janeiro, this suggests that there should already be many deaths that would not have been captured by conventional epidemiological surveillance system, based on individual notification of cases. COVID-19 is characterized by a majority of mild cases that can be confused clinically with influenza, or even with respiratory syncytial virus that occurs at this time of year in these two Brazilian cities[18]. Therefore, it is plausible to believe that SARS-CoV-2 has circulated widely and this circulation was not identified at the beginning of the Brazilian epidemic. As patients with COVID-19 worsen around the 10th day of symptom and death often occurs 3 weeks after the onset of symptoms, there may have been an important transmission even before the first case was identified. The County of Santa Clara Medical Examiner-Coroner identified SARS-CoV-2 at the autopsy of a patient who died at home because of an unknown cause on February 6 with no history of travel abroad. The first case of community transmission in the USA was identified by regular epidemiological surveillance on February 26 [19]. During the 2009 influenza pandemic in the State of São Paulo, it was possible to identify that mortality from respiratory diseases in the population was already above the upper limit of 95% of the expected two weeks before confirmation of community viral circulation in the State [13]. These findings reinforce the sensitivity and usefulness of surveillance of excess deaths and other complementary strategies of traditional epidemiological surveillance based on the notification and investigation of suspected clinical cases.

Another relevant aspect was the fact that the excess of mortality until week 16 had a strong Pearson correlation with official mortality (r = 0.94), this reinforces the consistency of the findings. The slope of the regression line indicates that the excess of deaths in the 6 cities was 2.5 times the number of deaths that were in the official surveillance system at the time. This demonstrates the potential of this type of surveillance in the opportunity to detect changes in the pattern of mortality, effectively and importantly complementing traditional epidemiological surveillance.

The present study has some limitations, many of which are a consequence of the fact that the assessment is made at the moment when the epidemic is still in its beginning, so some data may still be incomplete. In addition, death records were obtained from the registry information system instead of the SIM (Mortality Information System), which check the consistency of the diagnosis and follow the recommendations of the World Health Organization for the classification of the basic cause of death.. This choice was due to the fact that the SIM takes at least 6 months to consolidate the information and free access for consultations. We emphasize that this limitation is restricted to the analysis of the excess of deaths from respiratory causes, since the cause of death does not interfere in the assessment of mortality from all causes. Another limitation related to the use of data from the registry offices is that in this system only data are available for the years 2019 and 2020, therefore, it was not possible to use an average mortality rate of several years as would be the most appropriate [14,15]. Even with these limitations, the results were quite consistent, showing the most affected ages and there was a strong correlation between the mortality found in the official surveillance system and excess mortality, in addition, the period of increase in mortality was coincident with the pandemic. In addition, no excess deaths were identified in the city with a low incidence of COVID-19.

Excess mortality is a tool already used to assess mortality associated with climatic events, earthquakes and epidemics with good temporal definition such as those caused by the influenza virus, respiratory syncytial virus and chikungunya[13,14,20]. Like the CDC in the USA, the EuroMOMO Project tracks mortality data from some European countries weekly to assess excess mortality and monitor the impact of influenza epidemics, these systems are managing to capture the excess mortality associated with the COVID-19 pandemic. that is ongoing [15,21]. Obtaining excess deaths, regardless of the cause recorded on death certificates, seems to be the best estimate of mortality during the pandemic [22].

We believe that surveillance aimed at excess mortality is a simple, low-cost, easy-to-implement tool that can collaborate a lot with traditional epidemiological surveillance, therefore, it should have its use expanded to other countries and encouraged by the World Health Organization. Most countries already have mortality systems that could easily be adapted for this purpose. An important characteristic of this approach is that it does not depend on the flow of investigation of cases by health professionals, who are involved in preparedness and response activities during major epidemics. Because it does not necessarily depend on laboratory research, surveillance of excess mortality can be very useful, especially in low-income countries. In addition, it does not depend on the initial clinical suspicion, being able to more sensibly identify all deaths associated with the occurrence of different atypical phenomena, as diverse as epidemics, extreme weather events or earthquakes.

## Data Availability

This is a study based on official public data.
https://transparencia.registrocivil.org.br/registral-covid
https://sidra.ibge.gov.br/tabela/5918
https://coronavirus.saude.gov.br/

https://sidra.ibge.gov.br/tabela/5918

https://coronavirus.saude.gov.br/

https://brasil.io/dataset/covid19/caso/

